# Anticipating influenza-like illness outbreaks via syndromic surveillance using over-the-counter drug sales and primary health care data

**DOI:** 10.1101/2025.05.12.25327459

**Authors:** Juliane F. Oliveira, Thiago Cerqueira-Silva, Pedro Anibal N. Brito, Nívea B. da Silva, Rosemeire L. Fiaconne, Maria Célia S. L. Cunha, Gerson G. Cunha, Ray B. Miranda, Lucas M. Dias, Fábio M. H. S. Filho, Gerson O. Penna, Pablo Ivan P. Ramos, Izabel Marcilio, Manoel Barral-Netto, Viviane S. Boaventura

## Abstract

Effective pandemic preparedness relies on integrating diverse data sources for early outbreak detection. This study assessed whether over-the-counter (OTC) drug sales and primary health care (PHC) data could anticipate surges in respiratory-disease–related hospitalizations in Brazil. From November 2022 to June 2025, we analysed weekly time-series across 510 regions using a negative binomial autoregressive model within Statistical Process Control techniques. OTC data anticipated 56.6% of 746 hospitalization surges 1–3 weeks in advance, detected 9.5% concurrently, and missed 33.9%. PHC data anticipated 59.5%, detected 10.3% concurrently, and missed 30.2%. PHC data showed higher sensitivity and specificity than OTC (69.8% vs. 66.1%, and 49.5% vs. 47.8%). Performance varied regionally, and in 76.7% of regions, at least one stream showed high precision. These findings support the value of OTC drug sales, alongside PHC data, as early indicators of hospitalization surges in respiratory illness surveillance.

## Introduction

Early outbreak detection remains a critical challenge in public health^1–3^. Timely and accurate identification of unusual patterns that indicate the emergence or re-emergence of infectious diseases is essential for triggering rapid intervention, mitigating disease spread, and protecting vulnerable populations^4,5^. Traditional health surveillance refers to the systematic collection, analysis, and interpretation of structured health data, usually relying on predefined case definitions, for monitoring trends and estimating disease burden^6^. However, traditional surveillance methods generally present with timeliness and coverage limitations, and do not prioritize early warning^7^. To address this gap, syndromic surveillance is being increasingly used for early outbreak detection^2,8^, enabling the joint use of traditional surveillance sources with alternative, health-related data streams collected and analysed in near- or real-time^9^. In this context, data from over-the-counter (OTC) drug sales and administrative data of primary health care (PHC) encounters have been listed among the data streams with reported capabilities to improve outbreak detection^10–13^.

OTC drug sales data offer an accessible and real-time alternative for syndromic surveillance. They are widely available across countries, require minimal integration into the formal health systems, and capture consumer behaviour that often precedes health-seeking in formal medical settings^14^. Despite these attributes, OTC drug sales have been underused for early outbreak detection. Previous studies, mainly conducted in high-income countries ^15–19^, confirm the feasibility of using drug sales to identify respiratory disease outbreaks. However, the majority were published before 2015, and few have conducted real-time or prospective evaluations of OTC data. Key limitations include short observation periods, less than two years in most cases, which are not sufficient to assess seasonal patterns of disease outbreaks. Other reported limitations involve the absence of standardized drug classification systems, and restricted population or pharmacy coverage. Furthermore, few studies have systematically compared OTC data with other real-world, pre-hospital syndromic data sources such as PHC encounters.

Brazil presents a compelling case for advancing multi-source syndromic surveillance. As one of the world’s largest and most populous countries, Brazil faces substantial heterogeneity in disease dynamics, healthcare access, and data infrastructure. The country’s public Unified Health System (SUS) provides universal coverage while progressively incorporating digital health reporting. For instance, the recording of every publicly funded PHC encounter – that includes the date, municipality, and reason of encounter – is mandatory for resource allocation, resulting in a comprehensive and consistent national PHC information dataset^20^. In parallel, Brazil’s extensive private pharmacy network, comprising over 100,000 stores nationwide, generates high-frequency OTC drug sales data across a broad range of geographic and demographic contexts, which can be leveraged for health surveillance in complement to the data collected at the primary care level^21^.

This study leverages this unique setting to evaluate the potential of OTC drug sales and PHC encounter data for early detection of respiratory diseases-related hospitalization surges. We aim to: (i) detect warnings in each data stream using robust time-series modelling and anomaly detection methods; (ii) assess their capabilities to anticipate hospitalizations surges; and (iii) explore contextual factors, such as geographic characteristics, that may influence the early warning performance of these data streams.

## Results

### Data overview and descriptive analysis

Between November 2022 and June 2025, Brazil recorded 62,289,087 PHC encounters associated with ILI symptoms and 2,294,329 respiratory diseases-related hospitalizations, averaging 458,008 (95% CI: 432,605 – 483,411) PHC encounters and 16,870 (95% CI: 16,155 – 17,585) hospitalizations per week. Simultaneously, monitored pharmacies sold 689,297,990 drug units associated with treatment of ILI symptoms, with a mean weekly sale of 5,068,368 units (95% CI: 4,792,123 – 5,344,612).

At the national level, the number of PHC encounters, OTC drug sales and hospital admissions showed no significant trends in the study period. However, significant seasonal patterns were observed and maintained over the years, with all three data streams rising in March (epidemiological week [EW] 10). Records of PHC encounters and hospitalizations peaked in May (from EW 18 to 22), while OTC drug sales peaked later in July (from EW 27 to 31) (Supplementary Figure 2). However, when analysing trend and seasonality by immediate regions, distinct dynamics emerged based on the presence or absence of trends and seasonality (Supplementary Table 1).

Lagged cross-correlation analysis revealed strong temporal associations among surveillance streams across Brazilian immediate regions. Moderate to very strong correlations were observed between PHC encounters and OTC drug sales (78.6% of regions), PHC encounters and hospitalizations (94.7%), and OTC drug sales and hospitalizations (63.9%). For PHC-hospitalization pairs, 35.7% of regions showed synchronous correlations (lag 0), while 11.4% exhibited PHC encounters preceding hospitalizations by 1–3 weeks, while 47.7% showed the reverse pattern. Among OTC-hospitalization pairs, 16.3% were synchronous (lag 0), with OTC drug sales preceding hospitalizations by 1–3 weeks in 32.0% of regions and hospitalizations preceding OTC sales in 15.6%. PHC-OTC correlations were synchronous in 22.2% of regions, with OTC drug sales leading PHC encounters in 47.1% and PHC encounters leading OTC sales in 9.5%, both considering 1 to 3 weeks anticipation (Supplementary Table 2).

Lastly, to compare the spatial distribution of these counts across immediate regions, Figure 1 shows the number of cases per 100,000 inhabitants for PHC encounters, OTC drug sales, and hospitalizations over the study period (Supplementary Table 3).

**Figure 1:**
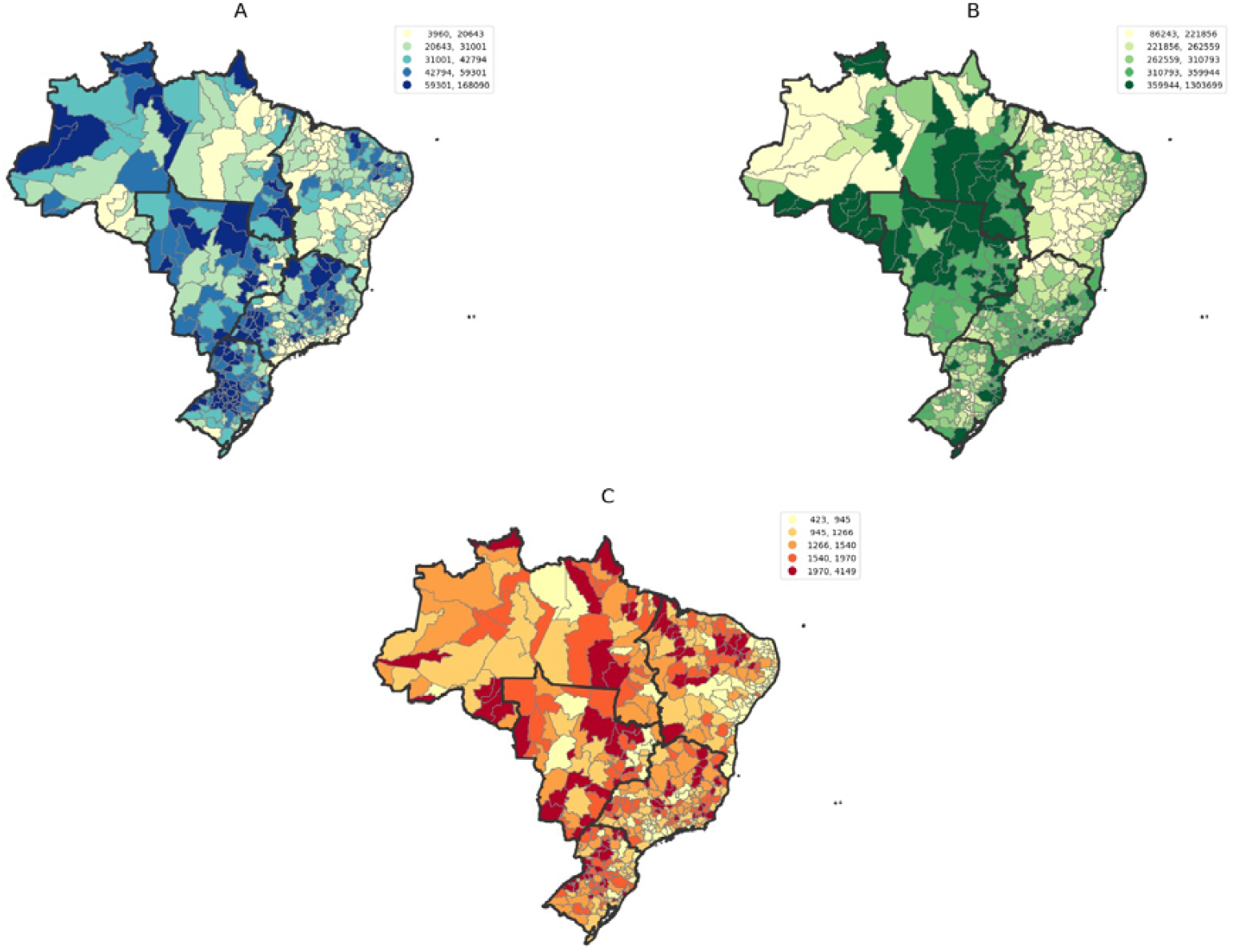
Spatial distribution of (A) ILI-related PHC encounters, (B) OTC drug units sold and (C) respiratory diseases-related hospitalizations across Brazil’s immediate regions from November 2022 to June 2025. To enhance comparability, the indicators are presented as rates per 100,000 inhabitants.

There were strong to very strong correlations between regional rates of PHC encounters (ρ = 0.704, p < 0.001) and OTC drug sales (ρ = 0.81, p < 0.001) with hospitalizations. Correlations were particularly high in the South and Center-West regions. For PHC encounters, the correlations reached ρ = 0.836 (p < 0.001) in the South and ρ = 0.810 (p < 0.001) in the Center-West. Similarly, for OTC drug sales, the correlations were ρ = 0.873 (p < 0.001) and ρ = 0.809 (p < 0.001), respectively. OTC drug sales also showed very strong correlation in the North (ρ = 0.869, p < 0.001), whereas PHC encounters exhibited the weakest correlation in that region (ρ = 0.618, p < 0.001) (Supplementary Table 4).

Overall, we observed a strong correlation between PHC encounters and OTC drug sales (ρ = 0.646, p-value < 0.001), with exception of the Southeast regions that showed a moderate correlation (ρ = 0.462, p-value < 0.001).

### Performance of anomaly detection

In the study period, 746 respiratory diseases-related hospitalization surges were identified in 469 (92.0%) of 510 immediate regions, of which 41 (8.0%) showed no hospitalization surges. Accordingly, by applying the NegBi-SPC model to each time series, we identified 13,106 warnings in PHC encounters and 13,066 in OTC drug sales. Of these, 10,442 (PHC) and 10,282 (OTC) occurred in regions that experienced at least one hospitalization surge (Supplementary Note 3 and Supplementary Data).

Warnings/surges emerged in February (EW 7) for PHC encounters and OTC sales, and in March (EW 10) for hospitalizations, aligning with expected nationwide seasonal patterns, as shown before. Notably, we also observed an increase in warnings starting in August (EW 32) for PHC encounters, followed by a similar pattern in OTC drug sales in September (EW 36) (Figure 2, Supplementary Table 5). However, we identified only five hospitalizations surges from August to December of 2023 and 12 surges from August to December of 2024.

**Figure 2:**
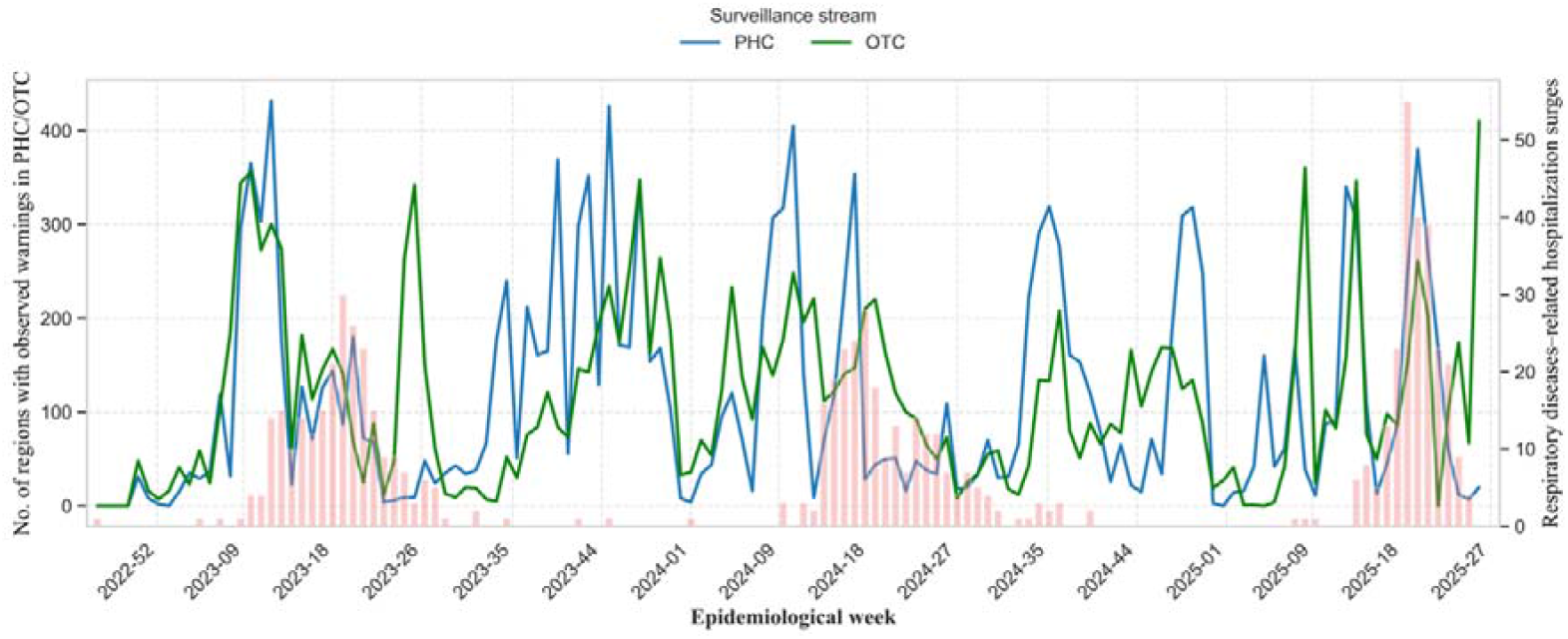
Number of regions with observed warnings in the PHC and OTC time series and/or hospitalization surges, per week, from November 2022 to June 2025. Counts of immediate regions with hospitalization surges in red, and warnings in OTC (green) and PHC (blue).

Figure 3 highlights the comparative timeliness performance of PHC and OTC data in anticipating hospitalization surges. Assessing the OTC time series, we could detect 56.6% (422/746) of hospitalization warnings 1 to 3 weeks in advance, compared with 59.5% (444/746) detected by the PHC encounters data. This represents 22 additional hospitalization surges anticipated by monitoring PHC series in relation to OTC alone. In addition, the PHC series had a slightly higher detection proportion of hospitalization surges within the same week: 10.3% (77/746) versus 9.5% (71/746). Also, the proportion of missed hospitalization surges was lower for PHC (30.2% (225/746)) than for OTC (33.9% (253/746)).

**Figure 3:**
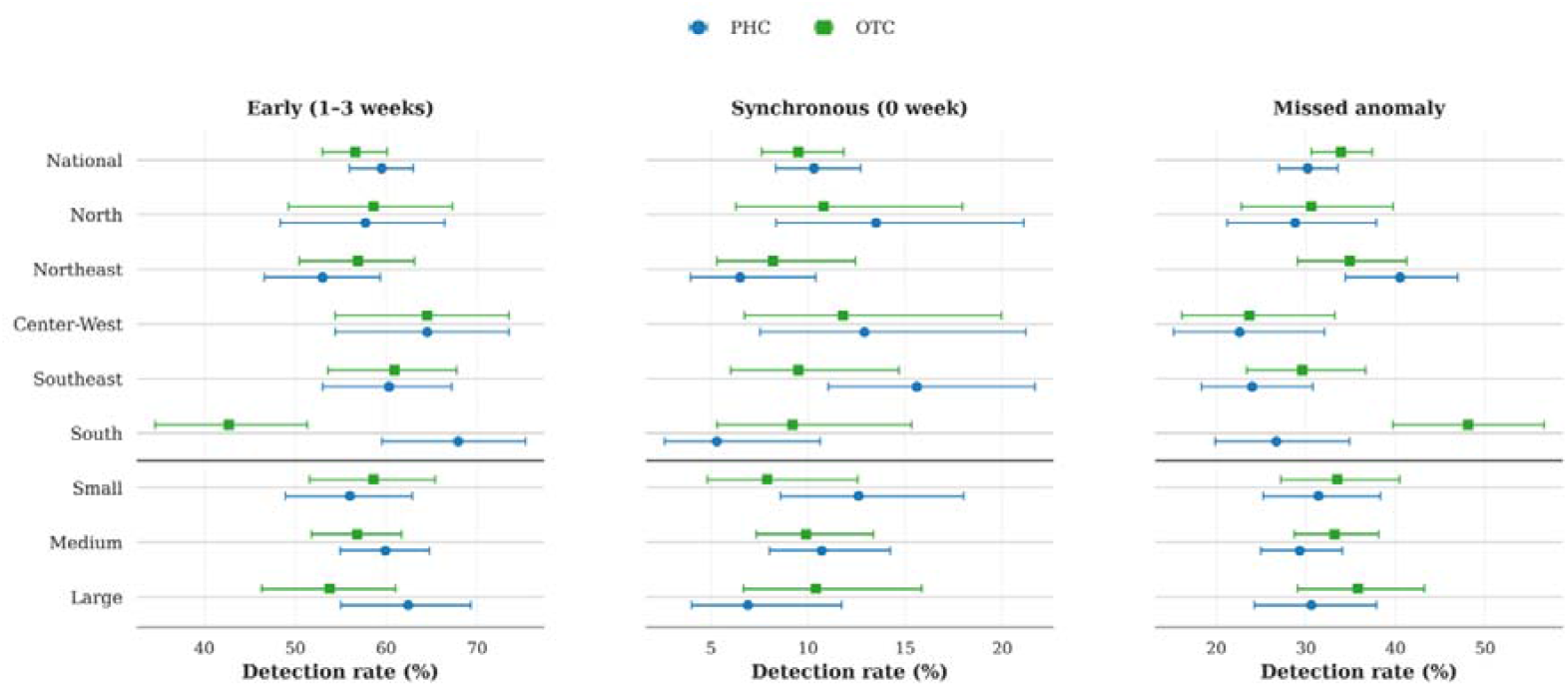
Timeliness of early warnings detected in PHC and OTC time series in relation to hospitalization surges at the national and regional levels. This figure summarizes the proportion of hospitalization surges anticipated (1 to 3 weeks early), detected in the same week (synchronous), or missed based on PHC and OTC series, along with their corresponding 95% confidence intervals (CIs). Results are presented for the national level, geographic region, and by population size.

The timeliness of PHC-based warnings varied across regions and population size. The Center-West region exhibited the highest early detection rates, with 64.5% (60/93) of warnings issued 1 to 3 weeks in advance to hospitalization surges. In contrast, the Northeast had slightly lower PHC timeliness, with 53.0% (123/232) issued 1 to 3 weeks in advance and 40.5% (94/232) of hospitalizations surges missed. Performance also varied by population size, with optimal early detection (1–3 weeks anticipation) in large-sized regions 62.4% (108/173) while small population regions showed reduced performance, with early anticipation rates of 56.0% (107/191) for PHC data. For OTC-based warnings the differences across regions between municipalities with different population sizes were less than 5% (Figure 4), with exception of South and large-sized regions.

**Figure 4:**
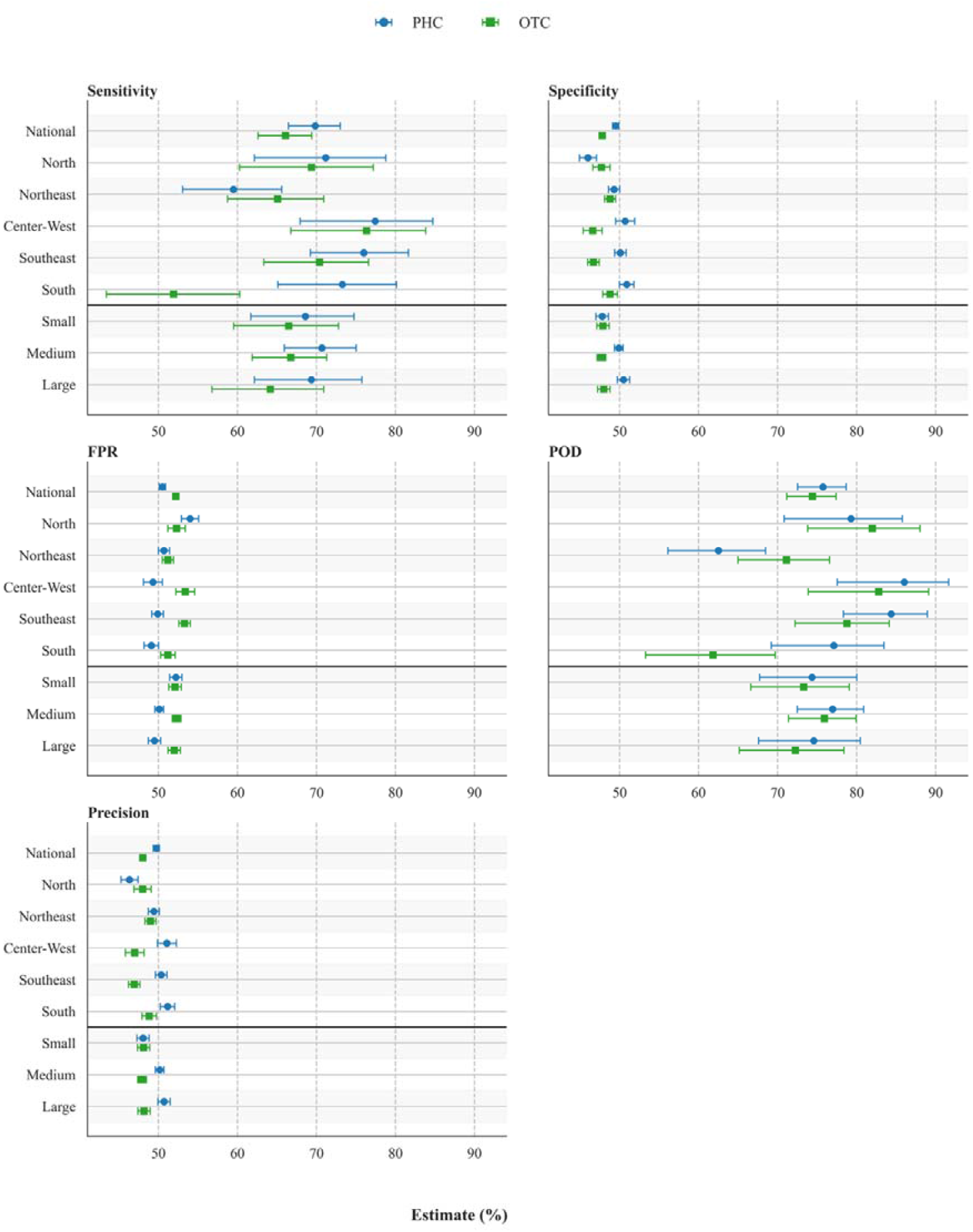
Performance metrics for PHC encounters and OTC drug sales data in detecting hospitalization surges across regions and population sizes. The figure presents sensitivity, specificity, probability of detection (POD), false positive rate (FPR), and precision for both PHC and OTC time series, along with their corresponding 95% CIs.

By assessing sensitivity and probability of detection, we verified that PHC encounters data (Se = 69.8% and POD = 75.7%) slightly outperformed OTC drug sales data (Se = 66.1% and POD = 74.4%), indicating improved early warning capabilities during the study period. OTC data also showed a higher false positive rate (FPR = 52.2% for OTC vs FPR = 50.5% for PHC) and lower precision (Pr = 48.0% for OTC vs Pr = 49.8% for PHC) across all regions, suggesting potential over-detection (Figure 4).

Both PHC encounters and OTC drug sales data streams exhibited better performance in the Center- West, achieving the highest Se (77.4% vs 76.3%) and POD (86.0% vs 82.8%) among all regions. Overall, PHC data generally outperformed OTC across most regions during the study period, with sensitivity ranging from 59.5% (Northeast) to 77.4% (Center-West) and POD between 62.5% (Northeast) to 86.0% (Center-West). The Northeast (Se = 59.5% and POD = 62.5% for PHC) and the South (Se = 51.9% and POD = 61.8% for OTC) showed the weakest performance for the PHC and OTC streams, respectively.

Regarding population size, medium-sized cities achieved the best overall performance for both PHC and OTC (Se = 70.7% for PHC, Se = 66.8% for OTC). In contrast, PHC data showed lower performance in small population regions (Se = 68.6% and FPR = 52.2%), while OTC performance declined in large population areas (Se = 64.2% and FPR = 52.0%).

### Complementary value of using OTC and PHC

Overall, 143 (28.0%) out of 510 regions demonstrated high precision in both PHC (Pr ≥ 50.0%) and OTC (Pr ≥ 47.8%) data streams. OTC data alone outperformed PHC in 125 (24.5%) regions, while

PHC performed better in 123 (24.1%) regions. In contrast, both data sources exhibited low precision or no data in 119 (23.3%) regions.

Regarding the timeliness score, both PHC and OTC data showed high anticipation in 202 (43.1%) regions. PHC outperformed OTC in 101 (21.5%) regions, whereas OTC had higher timeliness in 74 (15.8%) regions. In 133 (28.4%) regions, both data sources showed low anticipation or no data. Due to the absence of hospitalization surges, precision and timeliness could not be evaluated in 41 (8.0%) regions, represented in our results as no dada category (Figure 5).

**Figure 5:**
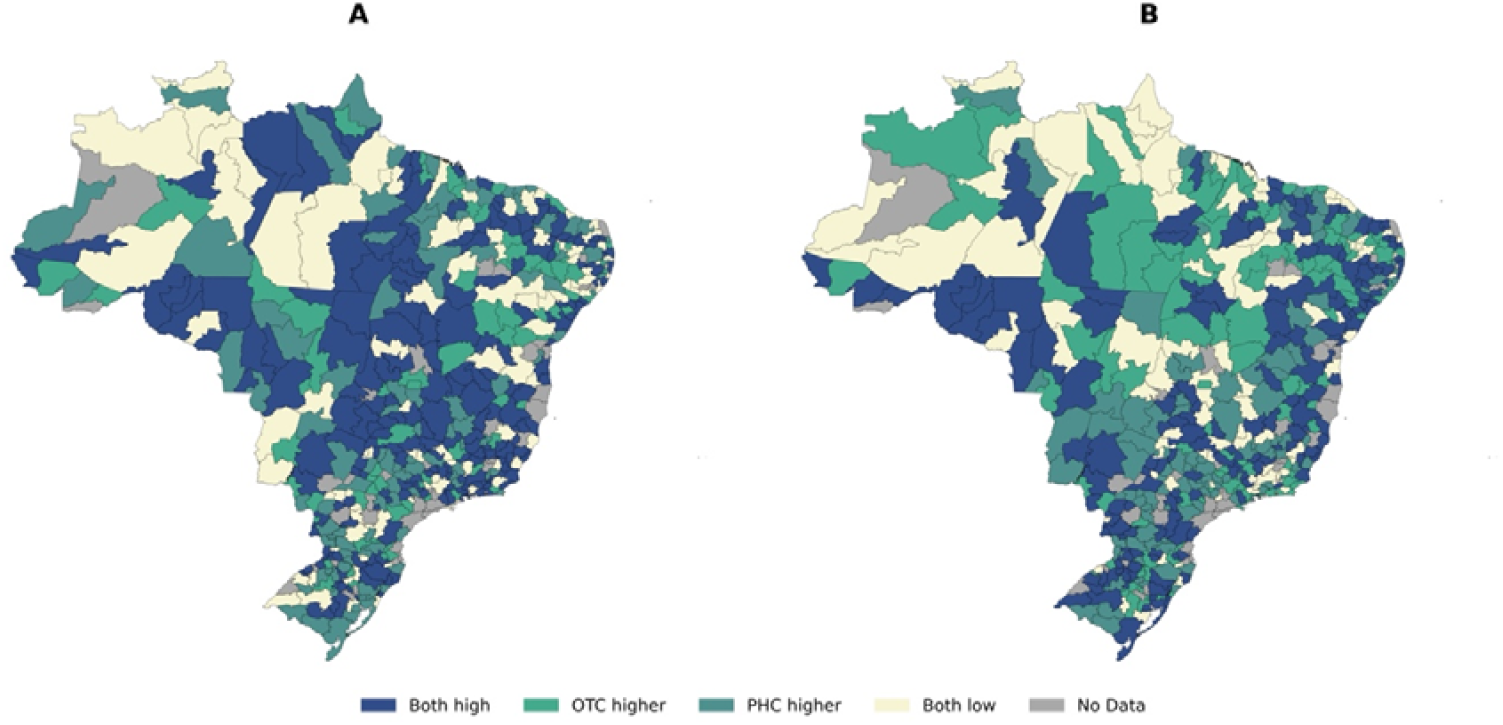
Classification of regions based on A) anticipation and B) precision indicators using PHC encounter and OTC drug sales data. A region was classified as “High” in anticipation, if anticipation scores were ≥ 60% for both PHC encounter and OTC drug sales data. For precision, regions with values ≥ 50.0% for PHC and ≥ 47.8% for OTC (median thresholds) were also considered “High”.

### Effect of alternative anomaly definition in hospitalizations time-series

We assessed the robustness of our findings by modifying the threshold criteria used to define hospitalization surges (Supplementary Note 2). Overall, the findings remain broadly consistent under the new definition. However, some shifts are observed, particularly in the timeliness patterns by population size, and in performance metrics when analysed by both macro-regions and population strata. Notably, the fundamental value of using PHC and OTC data for outbreak anticipation, individually or in conjunction, persisted across threshold definitions.

## Discussion

This study provides novel evidence that ILI-associated OTC drug sales can serve as early indicators of surges of respiratory diseases-related hospitalizations, providing 1–3 weeks of advance warning. We also observed that integrating ILI-associated OTC sales and PHC encounter data enhances the system’s capacity to anticipate these hospitalization surges. The use of OTC and PHC data show a good sensitivity performance and provide early detection of respiratory diseases-related hospitalization surges, but at the cost of increased false alarms. The core conclusion that PHC and OTC data are valuable for anticipating outbreaks held true regardless of how the threshold of hospitalization surges was defined. This complementary use of data streams with distinct operational strengths enables balanced surveillance systems capable of both timeliness detection and reliable warning, supporting more effective public health responses to emerging respiratory outbreaks.

While syndromic surveillance using multiple data sources with varied methodologies may enhance ILI outbreak trend detection, few studies have directly compared their effectiveness. In France, previous research suggested that drug sales were more sensitive and provided timelier signals than sentinel surveillance systems^17^. The high performance of OTC sales data observed in our analysis in a large, continental country like Brazil supports its potential use in areas where PHC data are unavailable. In these two countries OTC sales data proved highly effective as an independent surveillance tool. Previous studies in high-income countries also supported the utility of OTC drug sales for syndromic surveillance while our results extend this evidence to a low- and middle-income countries (LMIC) context^2,16,22–24^. As OTC data are routinely collected for commercial purposes, they usually provide high coverage of pharmaceutical sales, offering a representative proxy of medication consumption nationwide and creating no additional burden on the health system^2,9^. This is especially vital for rapidly extending surveillance in regions with limited public health infrastructure. These areas, often disproportionately affected by respiratory disease outbreaks due to fragile health systems, could benefit significantly from an early-warning system that requires no additional data collection burden. Using OTC sale for syndromic surveillance offers scalable, cost-effective solutions aligned with WHO pandemic preparedness goals, as set forth by the recently agreed proposal for a Pandemic Agreement^25^. Integrating such information from multiple countries could bridge surveillance gaps, enhancing early detection and targeted responses for respiratory infections and other communicable diseases.

Our findings indicate strong spatial and temporal alignment between OTC drug sales, PHC encounters, and hospitalizations, confirming that these routinely collected data sources capture epidemiologically meaningful disease fluctuations in Brazil. The warnings in PHC and OTC data also captured regular increases in ILI activity not associated with rise in hospitalizations. While non-confirmatory, concurrent increases in both medical visit patterns and self-medication behaviour suggests the occurrence of real, albeit mild, outbreaks. These increases in warnings from PHC encounters, which are sometimes preceded or followed by rises in warnings from OTC drug sales, reflect the fact that ILI is predominantly caused by self-limiting infections commonly managed with OTC medications. As a result, individuals often self-manage symptoms using OTC drugs and, consequently, do not seek additional care^26^. Detecting these early and less severe events is also important for guiding public health actions, as they may still place significant demand on primary care and serve as early indicators of emerging threats^27,28^. Interestingly, during out-of-season surges of respiratory diseases-related hospitalization periods, PHC warnings tend to precede those from OTC data, suggesting that OTC data are more likely to anticipate the onset of severe cases rather than the earliest signals of mild illness in the community. These findings underscore the need to evaluate the surveillance potential of these indicators using additional data sources, such as emergency unit admissions, to fully characterize the system’s performance across the disease severity spectrum. Finally, we also observe spikes in OTC warnings following hospitalization surges. One explanation for these spikes is increased drug consumption driven by promotional campaigns carried out by the pharmaceutical industry ^29^.

PHC and OTC data showed variation across Brazilian regions and by population strata, particularly in terms of timelines, sensitivity and probability of detection. This variability implies that the effectiveness of PHC and OTC data for early outbreak detection may require locally-referred calibration to ensure consistent performance. This finding is in alignment with our previous study^11^, which suggested the performance of PHC-based surveillance is context-dependent, potentially influenced by disparities in health system access and reporting practices across regions of different sizes. However, for both OTC and PHC data, the POD was lower in more populous regions. This is probable because in these urban settings early signs of illness may be diluted making it more difficult to detect surges^12^. These results suggest the importance of enhancing data granularity to improve early detection performance.

In practice, daily decision-making requires clarity on which data stream (PHC or OTC) offers more reliable early warning signals. The only study to directly explore this question, conducted by Najmi et al.^30^, suggests that analysing each data stream independently performs better for detecting early signs of ILI than combining both in a unified bivariate model. Here, we derived region-specific indicators that assess both timeliness and precision to guide local health managers in tailoring surveillance strategies. Our findings highlight Brazil’s highly heterogeneous landscape, where some regions benefit from both data sources, while others display complementary strengths.

Our study has strengths and limitations. More than 60% of surges in respiratory diseases-related hospitalizations were detected using PHC and OTC data—remarkable results considering the use of real-world, highly volatile time series in a setting without an integrated surveillance system. This performance was achieved through a national, integrative analysis in a continentally-sized LMIC, using routinely collected data with broad geographic coverage. However, while hospitalizations were used as the gold standard, this may underestimate outbreaks caused by highly transmissible but less severe pathogens. We were also unable to evaluate the performance of PHC and OTC data in 8.0% of regions due to the absence of hospitalization surges indicative of outbreak periods. A more comprehensive evaluation requires standardized, granular outbreak definitions from health authorities.

In conclusion, this work demonstrated that integrating OTC sales with primary□care encounters sharpens early□warning capability. These findings support the use of OTC data as a robust, scalable, low-burden alternative for strengthening early warning systems in resource-limited settings and for broader application across countries aiming to improve epidemic preparedness. Longer series and pathogen□specific analyses (e.g., arboviruses, diarrhoeal diseases) will clarify how generalisable this approach is beyond respiratory syndromes.

## Methods

### Data sources

We analysed weekly time series data on OTC drug sales and PHC encounters, aggregated across 510 immediate regions in Brazil. Each region represents groupings of municipalities strongly connected to local urban centres, serving population needs for goods, employment, health services, and education (Supplementary Figure 1), as defined by the Brazilian Institute of Geography and Statistics (IBGE). The dataset covers the period from November 2022 to June 2025.

We used OTC sales data from IQVIA, a global health data and technology company whose coverage exceeds 95% of the Brazilian pharmaceutical retail market, providing a robust source for analysing medication purchasing trends^21,31^. The data includes the weekly number of medication units sold, covering 77 active substances and 54 drug classes categorized according to the Anatomical Therapeutic Chemical (ATC4) classification system. We filtered the data to include only drug classes frequently used for Influenza-like illness (ILI) treatment and applied a cluster analysis to group medications with similar temporal patterns, in accordance with known ILI OTC drug sales peaks in state capitals. Based on this analysis, six drugs were selected for surveillance: Ascorbic Acid, Chlorphenamine Maleate, Phenylephrine, Paracetamol, Loratadine, and Benzydamine Hydrochloride (Supplementary Note 1).

PHC data were extracted from the Brazilian Health Information System for Primary Care (SISAB), a database that registers data on all publicly-funded PHC encounters in Brazil, coded by either the International Classification of Diseases (ICD-10) or the International Classification of Primary Care (ICPC-2). ILI related encounters were filtered to construct an ILI-specific time series following the approach of Cerqueira-Silva et al ^11^.

To assess the ability of these series to anticipate the burden of respiratory diseases-related hospitalizations, we extracted hospitalizations cases, as per the ICD-10 (Chapter X - Diseases of the respiratory system, code range J00-J99) from the Brazilian Hospital Information System (SIH), covering the period from November 2022 to June 2025. Hospitalization data was used as the gold standard for evaluating the performance of each data stream.

We also used 2024 IBGE population estimates to classify regions by population size: small (≤ 117,789 inhabitants), medium (117,790 – 324,768 inhabitants), and large (> 324,768 inhabitants).

### Statistical Model

To evaluate the performance of OTC drug sales and PHC encounter data in providing early warnings of surges in respiratory disease–related hospitalization, we employed a series of negative binomial autoregressive models that best predict each time-series for each of the 510 immediate regions in Brazil. Negative binomial models are well suited for counting data that exhibit overdispersion, which is a key feature of our time series. Using the estimated values obtained by the best fitted model, we applied the Statistical Process Control technique to establish epidemic thresholds that are used to flag anomalous values in each time series^32,33^. We name the procedure as NegBi-SPC.

To define the NegBi-SPC procedure, let *y*_*t*_ for *t* = 1, …,*n*, denote the weekly (four windows) moving average of counts of OTC or PHC encounters, modeled as a negative binomial distribution. The NegBi-SPC model is defined in equation (1):

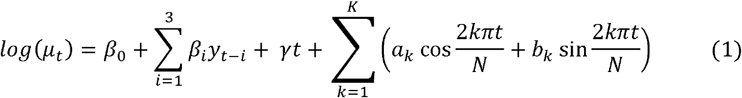

where *μ*_*t*_ is the mean at time t, *β*_0_ is the intercept, *β*_*i*_ are the coefficients for the autoregressive terms varying from *i* = 1 to 3. The coefficient *γ* captures the long-term trend in the series, indicating whether the mean level is increasing or decreasing over the study period. The summation of sine and cosine terms represents the seasonal component (a periodic pattern that repeats at regular intervals within the time series) with period N = 52 weeks, Fourier coefficients for the k-th harmonic, and K denotes the maximum number of harmonics included to adequately capture the seasonal structure of the time series.

Prior to including trend and seasonality terms, we first fit a negative binomial regression model including only the trend. We then assessed its statistical significance (p-value < 0.05) as a criterion for trend term inclusion in subsequent models. The seasonality component was evaluated using the Friedman test and included if found significant. When the test indicated significant seasonality, we applied the scipy.fft module to decompose the series and extract the relevant K Fourier coefficients for inclusion into the model. For each time series, we tested multiple model specifications, including combinations of trend, seasonality, and lagged terms. Models were estimated via maximum likelihood estimation, and the best-fitting model was selected based on the Akaike Information Criterion (AIC).

After model selection, lower control limits (LCL) and upper control limits (UCL) were calculated to flag anomalous values as described by equations (2) and (3):

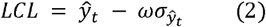

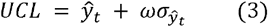

where 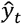 is the value estimated by the model at time 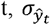 is the standard deviation estimate of the response predicted, and *ω* represents the distance between the control limits. Control limits is a statistical indicator on control charts to monitor the expected range of variation of the process. If data points fall outside the control limits or show unusual patterns, it signals that the process may need attention or adjustment. Throughout the text, we refer to anomalies found in OTC or PHC time series as warnings.

### Performances of early detection of respiratory diseases-related hospitalization surges

To evaluate the performance of early outbreak detection, anomalies in respiratory diseases-related hospitalizations were defined as weekly counts exceeding the historical median from November 2022 to June 2025, plus two standard deviations. An isolated anomaly was defined as a single week with an anomaly, which was not followed or preceded by any other anomaly within a period of three weeks. Isolated anomalies were excluded, as they are more likely to represent transient fluctuations than a consistent signal of an increase in hospitalization. On the other hand, consecutive anomalies were defined as two or more anomalous weeks occurring consecutively or separated by only one non-anomaly week. Consecutive anomalies were grouped together marking periods of sustained respiratory diseases-related hospitalization. The starting weeks of these periods are named as hospitalization surges. To assess the robustness of this definition, we added one more alternative criterion for identifying surges in the hospitalization time series in Supplementary Note 2.

We then assessed the performance of OTC and PHC time series in anticipating hospitalization surges using the following metrics^32^: Probability of Detection (POD), defined as the probability that a warning is generated at least once between the start and the end of an hospitalization surge; False Positive Rate (FPR), as the proportion of weeks corresponding to an alarm in the absence of a surge; Sensitivity (Se) and Specificity (Sp) are defined as Se = TP/(TP+FN) and Sp = TN/(TN+FP), where True Positives (TP) are warnings associated with hospitalization surge, True Negatives (TN) are weeks without warnings and hospitalization surges, False Negatives (FN) are weeks with hospitalization surge and without associated warnings, False Positives (FP) are weeks without hospitalization surges and with warnings. Lastly, we defined Precision (Pr) as Pr = (TP + TN) / (TP + TN + FP + FN). Immediate regions that did not present hospitalization surges during the study period were not included in this analysis.

Lastly, we conducted descriptive analyses to complement our main results. To explore how the proportions of PHC encounters, OTC drug sales, and hospitalizations vary across regions, we calculated Spearman correlation coefficients. Additionally, to examine the temporal relationships between each pair of time series within each immediate region, we computed lagged Spearman cross-correlations. The strength of correlation was classified as follows: very weak (ρ = 0.00–0.19), weak (ρ = 0.20–0.39), moderate (ρ = 0.40–0.59), strong (ρ = 0.60–0.79), and very strong (ρ = 0.80–1.00).

### Complementary value of using OTC and PHC

To assess the complementary value of OTC and PHC data sources for detecting hospitalization surges, we calculated two metrics for each immediate region: precision, as previously described; and a timeliness score, defined as the proportion of warnings occurring either during or up to three weeks prior to a hospitalization surge, relative to the total number of hospitalization surges in that region. A region was classified as having high precision if its precision value was above the median for all regions, and as having high timeliness performance if its timeliness score was higher than 60%.

## Data Availability

Under the terms of our data use agreements with the Brazilian Ministry of Health and IQVIA, we are not authorized to share the primary datasets with third parties. Researchers interested in accessing these data should submit requests directly to both parties.

## Acknowledgments

This study is part of the Alert-Early System of Outbreaks with Pandemic Potential (ÆSOP, http://aesop.health), an initiative under development by Brazil’s Fundação Oswaldo Cruz (Fiocruz) and the Federal University of Rio de Janeiro with financial support from the Rockefeller Foundation’s Health Initiative (Grant 2023-PPI-007 awarded to MB-N). MB-N, PIPR, and VB are CNPq Research Fellows. TC-S acknowledges funding from the Royal Society (NIF\R1\231435). The excellent project management and administrative support from Fundação de Apoio à Fiocruz (FIOTEC) is greatly appreciated. We acknowledge Meiruze Sousa Freitas for the strategic support provided during the development of this work. We acknowledge the great support of ACESSA (https://acessa.org.br/) and IQVIA for procuring and granting access to drug sales data.

## Data Availability

The data that support the findings of this study are available from the Brazilian Ministry of Health and IQVIA but restrictions apply to the availability of these data, which were used under licence for the current study, and so are not publicly available. Data are however available from the authors upon reasonable request and with permission of Brazilian Ministry of Health and IQVIA.

## Code Availability

All generated data and codes to ensure reproducibility of our results are available in our GitHub repository^34^.

## Author contributions

Conceptualization: JFO, MB-N, VB; Methodology: JFO, TC-S, PANB, NBS, RLF, FMHSF, VB; Investigation: JFO, TC-S, PANB, GOP, IM, PIPR; Visualization: JFO, MCSLC, GGC, RBM, LMD, PIPR; Funding acquisition: MB-N; Project administration: IM, MB-N, PIPR; Supervision: MB-N; Writing – original draft: JFO, VB; Writing – review & editing: all authors.

