## Supplementary File for "Anticipating influenza-like illness outbreaks via syndromic surveillance using over-the-counter drug sales and primary health care data"

#### The PDF file includes:

**Supplementary Figures 1 and 2**

Supplementary Figure 1: Map of Brazil's Immediate Geographic Regions

Supplementary Figure 2: Temporal trends of influenza-like illness indicators in Brazil and across Brazilian regions

**Supplementary Table 1 to 5**

Supplementary Table 1: Summary of immediate regions by presence of statistically significant trend and seasonality.

Supplementary Table 2: Lagged correlations between PHC encounters, respiratory diseases associated with hospitalizations, and OTC drug sales across immediate regions.

Supplementary Table 3: Total counts and rates (per 100,000 inhabitants) of respiratory diseases associated with hospitalization cases, ILI-related PHC encounters, and OTC drug units sold across Brazil and by region.

Supplementary Table 4: Monthly counts of respiratory diseases associated with hospitalizations surges and warnings in ILI-related PHC encounters and OTC drug sales.

Supplementary Table 5: Spearman correlation coefficients between respiratory diseases associated with hospitalization counts, PHC visits, and OTC drug sales at the national and regional levels.

**Supplementary Note 1 and 3**

Supplementary Note 1: OTC medications cluster analysis.

Supplementary Note 2: Alternative definitions for outbreaks in respiratory diseases associated with hospitalizations.

Supplementary Note 3: Model diagnostics

**Supplementary Data**

Data containing the variables supporting the evaluation of the NegBi-SPC model results.

**Supplementary references**

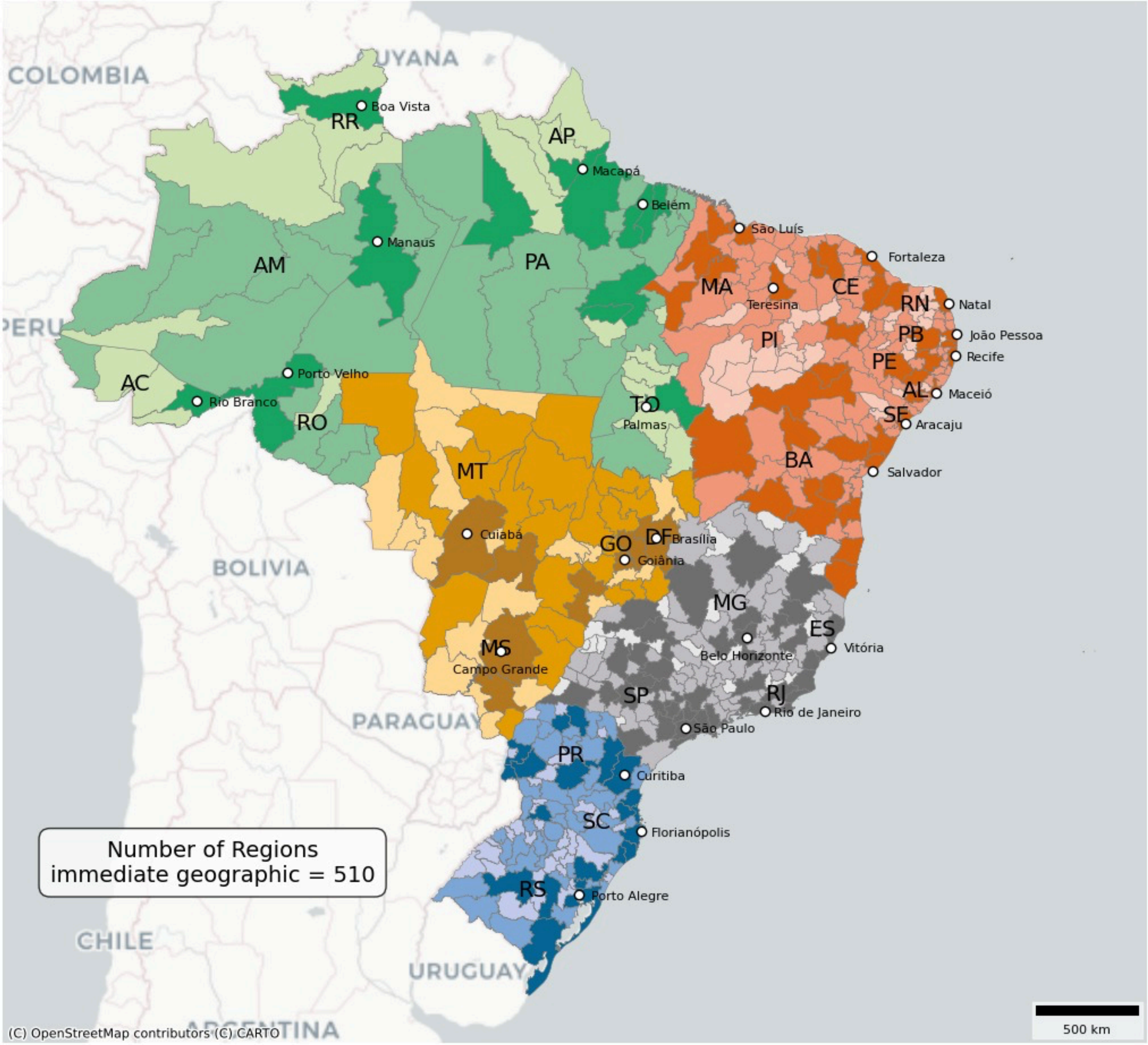


**Supplementary Figure 1**: **Map of Brazil's Immediate Geographic Regions, as defined by the Brazilian Institute of Geography and Statistics (IBGE).** The main colors represent Brazil's five major macro-regions: North (green), Northeast (red), Centre-West (orange), Southeast (grey), and South (blue). Within each macro-region, three shades of color indicate population strata of the immediate geographic regions: light for small populations (≤117,789 inhabitants), medium for populations between 5117,790 and 324,768 inhabitants, and dark for large populations (>324,768 inhabitants).

### Brazil

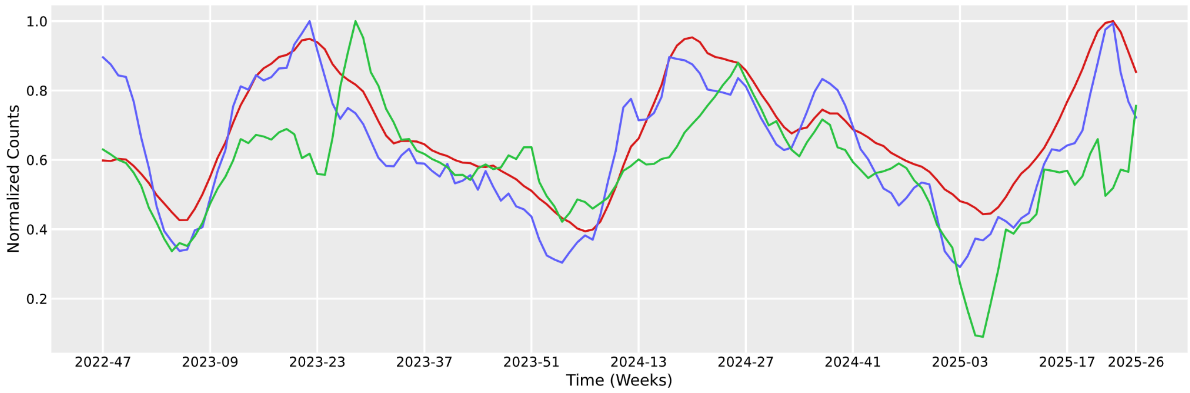


### North

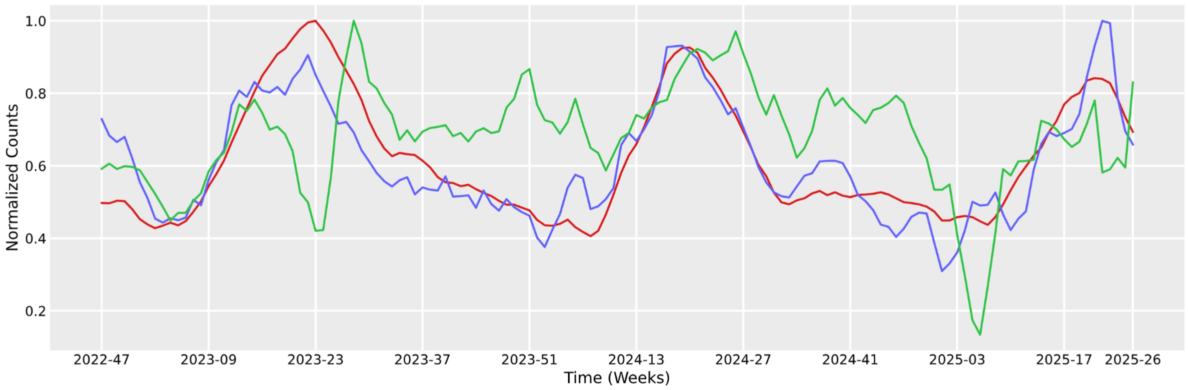


### Northeast

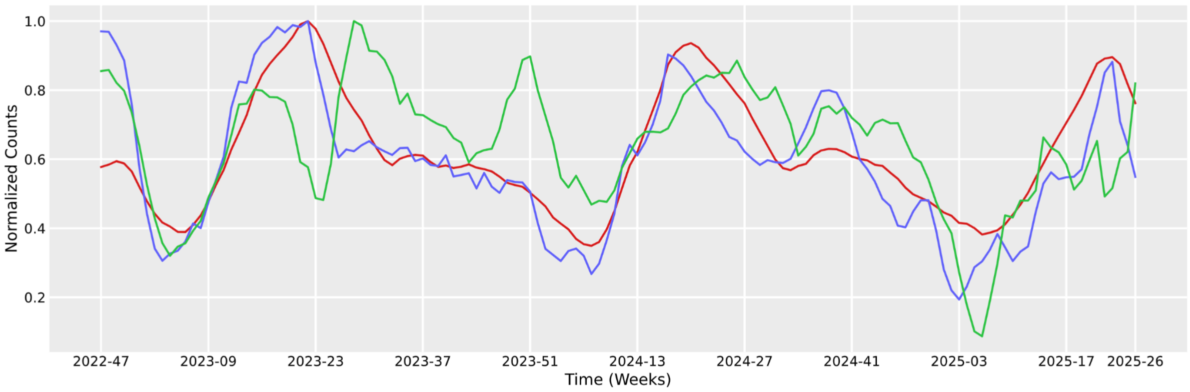

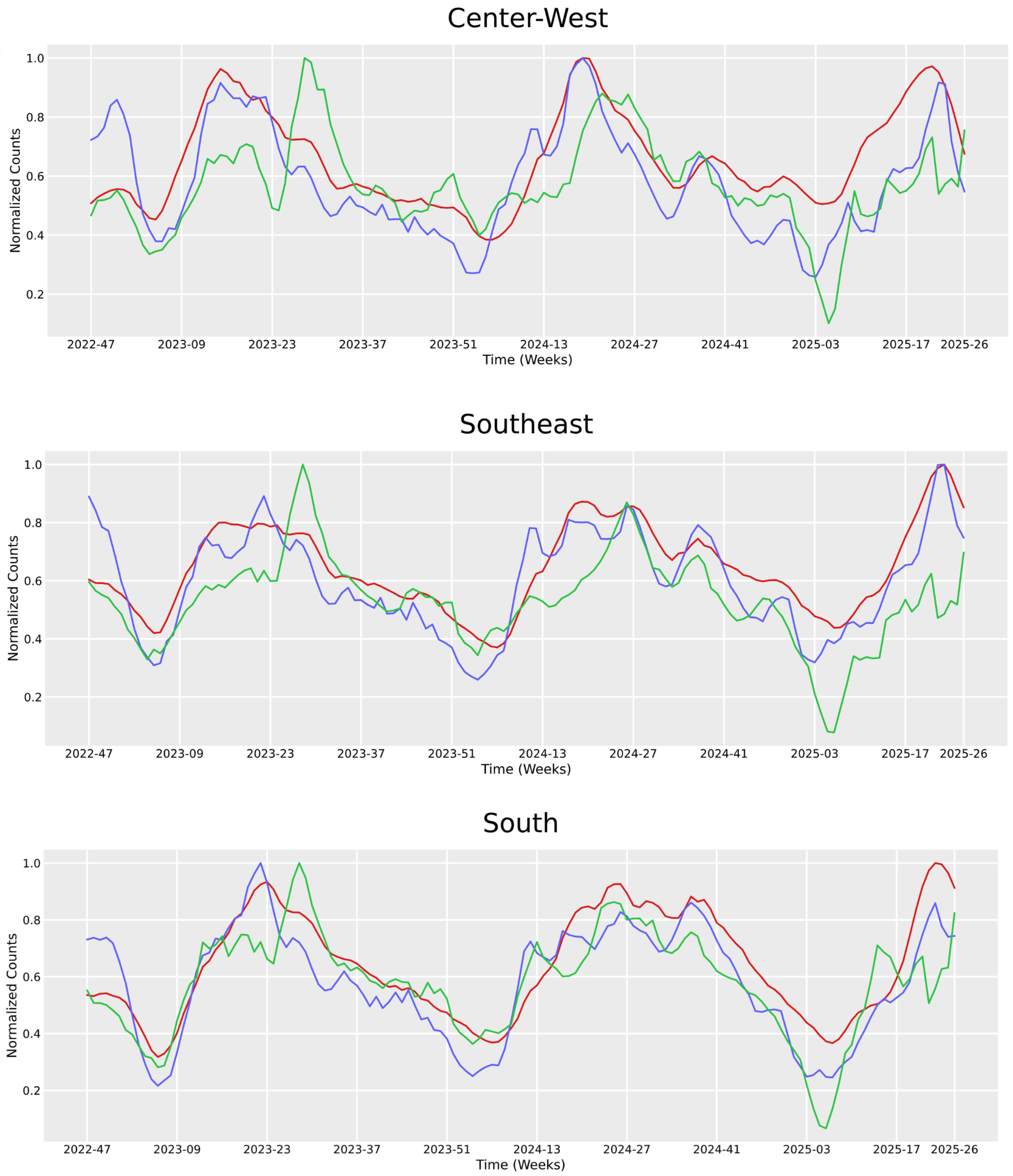


**Supplementary Figure 2**: **Temporal trends of influenza-like illness indicators for Brazil and across Brazilian regions.** Each subplot represents a different region, showing the normalized 4-week moving average of PHC encounters (blue), over-the-counter drug sales (green), and hospitalizations (red) from November 2022 to June 2025.

**Supplementary Table 1: Summary of immediate regions by presence of statistically significant trend and seasonality in time series of PHC Encounters, OTC Drug Sales, and Hospitalizations (Nov 2022–Jun 2025)**. Trend significance was determined using a negative binomial regression model, considering a p-value ≤ 0.05 for the trend term. Seasonality was assessed using the Friedman test.

| | Regions only with significant trend n (%) | Regions with significant trend and seasonality | Regions only with significant seasonality | Regions with no significant trend and seasonality |
|---|---|---|---|---|
| PHC encounters | - | 30 (5.9%) | 472 (92.5%) | 8 (1.6%) |
| Hospitalizations | 6 (1.2%) | 22 (4.3%) | 434 (85.1%) | 48 (9.4%) |
| OTC drug sales | 44 (8.6%) | 38 (7.5%) | 268 (52.5%) | 160 (31.4%) |

**Supplementary Table 2: Lagged correlations between PHC encounters, hospitalizations, and OTC drug sales across immediate regions.** The table shows the number and percentage of regions exhibiting moderate to very strong correlations at different lags. The last row represents the number of regions with weak or very weak correlations.

| **Lag** | **PHC encounters and hospitalizations n ( %)** | **OTC drug sales and hospitalizations n ( %)** | **PHC encounters and OTC drug sales n ( %)** |
|---|---|---|---|
| -3 | 5 (1.0) | 45 (10.8) | 87 (17.1) |
| -2 | 11 (2.2) | 49 (9.6) | 44 (8.6) |
| -1 | 42 (8.2) | 59 (11.6) | 109 (21.4) |
| 0 | 182 (35.7) | 83 (16.3) | 113 (22.2) |
| 1 | 189 (37.1) | 39 (7.6) | 35 (6.9) |
| 2 | 38 (7.5) | 19 (3.7) | 9 (1.8) |
| 3 | 16 (3.1) | 22 (4.3) | 4 (0.8) |
| **Weak or very weak** | 27 (5.3) | 184 (36.1) | 109 (21.4) |

**Supplementary Table 3: Total counts and rates (per 100,000 inhabitants) of hospitalization cases, ILI-related PHC encounters, and OTC drug units sold across Brazil and by region.** This table summarizes the total number of cases or units for each data source at the national level and stratified by geographic region (North, Northeast, South, Southeast, and Center-West). The values in parentheses represent the respective rates per 100,000 inhabitants in each region.

| **Data source** | **Total** | **North** | **Northeast** | **South** | **Southeast** | **Center-West** |
|---|---|---|---|---|---|---|
| Hospitalizations | 2,294,329 (1,129.9) | 238,934 (10.1) | 608,483 (8.2) | 396,635 (9.7) | 849,353 (7.4) | 200,924 (9.1) |
| ILI-related PHC encounters | 62,289,087 (30,674.8) | 5,190,804 (29,918.8) | 13,182,157 (24,123.4) | 14,159,036 (47,301.9) | 23,688,040 (27,918.5) | 6,069,050 (37,261.3) |
| ILI-related OTC drug units sold | 689,297,990 (339,451.1) | 67,963,092 (391,726.7) | 155,897,994 (285,294.5) | 96,350,138 (321,882.6) | 304,962,092 (359,425.1) | 64,124,674 (393,697.4) |

**Supplementary Table 4: Spearman correlation coefficients between hospitalization counts, PHC visits, and OTC drug sales at the national and regional levels.** This table presents the Spearman correlation coefficients and corresponding p-values for the relationships between hospitalization cases, PHC visits, and OTC drug sales across Brazil and within each geographic region (North, Northeast, South, Southeast, and Center-West). Correlation strength is classified as Very Weak (0.00–0.19), Weak (0.20–0.39), Moderate (0.40–0.59), Strong (0.60–0.79), and Very Strong (0.80–1.00).

| Region | Comparison | Spearman Correlation | P-Value | Strength |
|---|---|---|---|---|
| Country | Hospitalization and PHC visits | 0.704 | p-value < 0.001 | Strong |
| | Hospitalizations and OTC | 0.810 | p-value < 0.001 | Very Strong |
| | PHC visits and OTC | 0.646 | p-value < 0.001 | Strong |
| North | Hospitalization and PHC visits | 0.618292 | p-value < 0.001 | Strong |
| | Hospitalizations and OTC | 0.868903 | p-value < 0.001 | Very Strong |
| | PHC visits and OTC | 0.704465 | p-value < 0.001 | Strong |
| Northeast | Hospitalization and PHC visits | 0.734041 | p-value < 0.001 | Strong |
| | Hospitalizations and OTC | 0.748204 | p-value < 0.001 | Strong |
| | PHC visits and OTC | 0.791836 | p-value < 0.001 | Strong |
| Southeast | Hospitalization and PHC visits | 0.645386 | p-value < 0.001 | Strong |
| | Hospitalizations and OTC | 0.798699 | p-value < 0.001 | Strong |
| | PHC visits and OTC | 0.461648 | p-value < 0.001 | Moderate |
| South | Hospitalization and PHC visits | 0.835539 | p-value < 0.001 | Very Strong |

| | | | | |
|---|---|---|---|---|
| | Hospitalizations and OTC | 0.872625 | p-value < 0.001 | Very Strong |
| | PHC visits and OTC | 0.740640 | p-value < 0.001 | Strong |
| Center-West | Hospitalization and PHC visits | 0.810434 | p-value < 0.001 | Very Strong |
| | Hospitalizations and OTC | 0.808821 | p-value < 0.001 | Very Strong |
| | PHC visits and OTC | 0.786728 | p-value < 0.001 | Strong |

**Supplementary Table 5: Monthly counts of outbreak events (hospitalizations) and anomalies in ILI-related PHC encounters and OTC drug sales**, in regions with hospitalization warnings (Nov 2022–June 2025).

| **Month** | **Outbreak Events in Hospitalizations** | **Anomalies in ILI-related PHC Encounters** | **Anomalies in ILI-related OTC Drug Sales** |
|---|---|---|---|
| Total | 746 | 15,781 | 15,500 |
| January | 2 | 610 | 704 |
| February | 4 | 1,387 | 1,757 |
| March | 52 | 2,982 | 2,758 |
| April | 210 | 1,402 | 1,544 |
| May | 319 | 1,776 | 1,708 |
| June | 107 | 355 | 1,700 |
| July | 34 | 355 | 434 |
| August | 8 | 1,132 | 290 |
| September | 7 | 1,619 | 871 |
| October | 1 | 1,396 | 1,037 |
| November | 2 | 1,418 | 1,594 |
| December | 0 | 1,349 | 1,103 |

#### Supplementary Note 1

OTC medications cluster analysis

To identify OTC medications with similar sales dynamics and validate their utility as indicators of community-level respiratory disease activity, we performed a time series clustering analysis using the R package dtwclust (https://cran.r-project.org/web/packages/dtwclust/index.html). This analysis grouped drug sales time series based on their temporal behavior across Brazilian state capitals. We applied hierarchical clustering with the DIvisive ANAlysis (DIANA) method and used dynamic time warping (DTW) as the distance metric, allowing for flexible alignment of time series with local time shifts. A Sakoe-Chiba band with a window size of 8 weeks was imposed to constrain the alignment and avoid overfitting to noise.

Weekly sales data for each medication, aggregated at the state capital level from November 2022 to June 2025, were used as input time series. We explored clustering solutions ranging from 2 to 7 clusters and selected the optimal number based on the highest average Silhouette index, which evaluates cluster compactness and separation.

Drugs grouped in clusters whose temporal patterns exhibited strong alignment with full ILI OTC drug sales series were selected for further analysis. These clusters were considered most relevant for surveillance purposes, and the final selection of six drugs was informed by their presence in these clusters and their clinical relevance for treating influenza-like illness symptoms.

#### Supplementary Note 2

Alternative definitions for outbreaks  in hospitalizations due to respiratory diseases

To explore the performance of using ILI related PHC encounters and OTC drug sales data, we added here one more alternative criteria to define weekly respiratory diseases-related hospitalization surge. Hospitalization surges thresholds are defined based on the median weekly hospitalizations (p50) in the study period from November 2022 to June 2025. Specifically, regions are stratified by their historical median, and a corresponding threshold is applied, as summarised in the table below. To reduce noise in regions with very few hospitalizations (p50 < 50), a surge is only flagged if at least 10 hospitalizations occur in the week[1].

| Median (p50) of hospitalisations due to respiratory causes | Threshold |
|---|---|

| | |
|---|---|
| p50<50 | N > (2 * p50) & N>10 |
| 50 ≤ p50 <100 | N > (p50 + 0.5*p50) |
| 100 ≤ p50 <250 | N > (p50 + 0.4*p50) |
| 250 ≤ p50 <500 | N > (p50 + 0.3*p50) |
| 500 ≤ p50 <1000 | N > (p50 + 0.2*p50) |
| p50≥1000 | N > (p50 + 0.1*p50) |

Using the revised surge definition described above, we identified a total of 642 respiratory diseases-related hospitalization surges, occurring in 343 (67.3%) immediate regions. Compared to the anomaly-based definition used in the main manuscript, this new approach results in 126 fewer regions issuing surge events and identifies a larger number of regions (167; 32.7 %) that do not meet the criteria to trigger a surge in the hospitalization time series. Accordingly, restricting the analysis to only the regions that experienced at least one hospitalization outbreak event during the study period, we detected 11,700 anomalies in ILI-related PHC encounter time series and 11,308 anomalies in OTC drug sales data.

Despite the change in definition, the temporal dynamics of weekly counts of immediate regions issuing hospitalization outbreak events remain consistent (see figure 1 below).

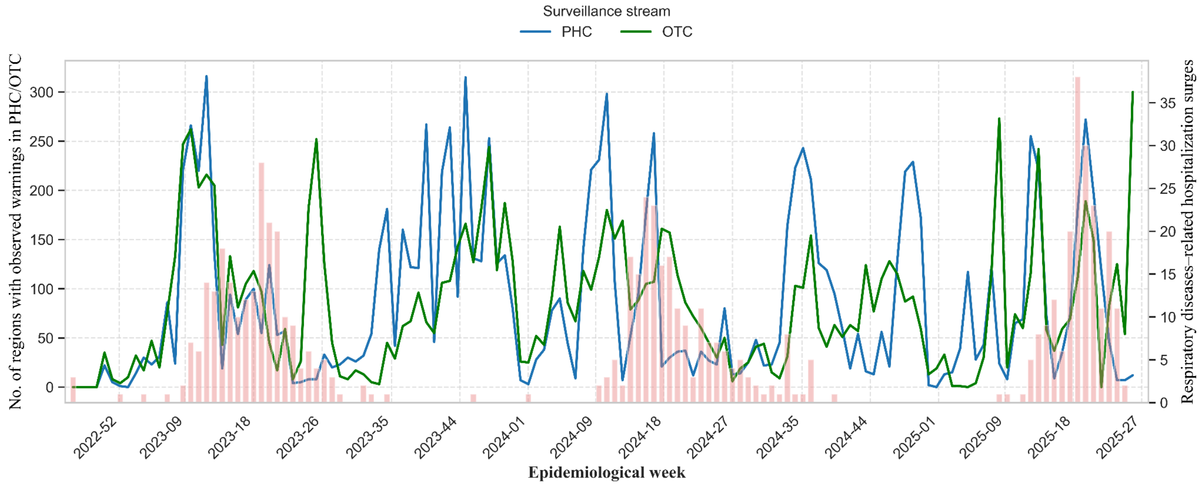


**Figure 1:** Weekly count of immediate regions in Brazil issuing warnings in PHC and OTC from November 2022 to June 2025, based on the revised definition. Counts of hospitalization surges in red, and counts of warnings in OTC in blue and PHC in green.

While the timeliness patterns of PHC and OTC data remain broadly consistent under the new definition, some shifts are observed in the stratification by population size (Figure 2 below). In contrast to the main manuscript, where both PHC and OTC data showed the highest proportion of early warnings (1 to 3 weeks in advance) in large and small-sized regions, respectively, this analysis finds the best anticipation occurring in small-sized regions, with lower proportions in large regions for PHC, and best anticipation in large-sized and lower in medium regions for OTC. The proportion of missed outbreak events also differs from the results reported in the main manuscript.

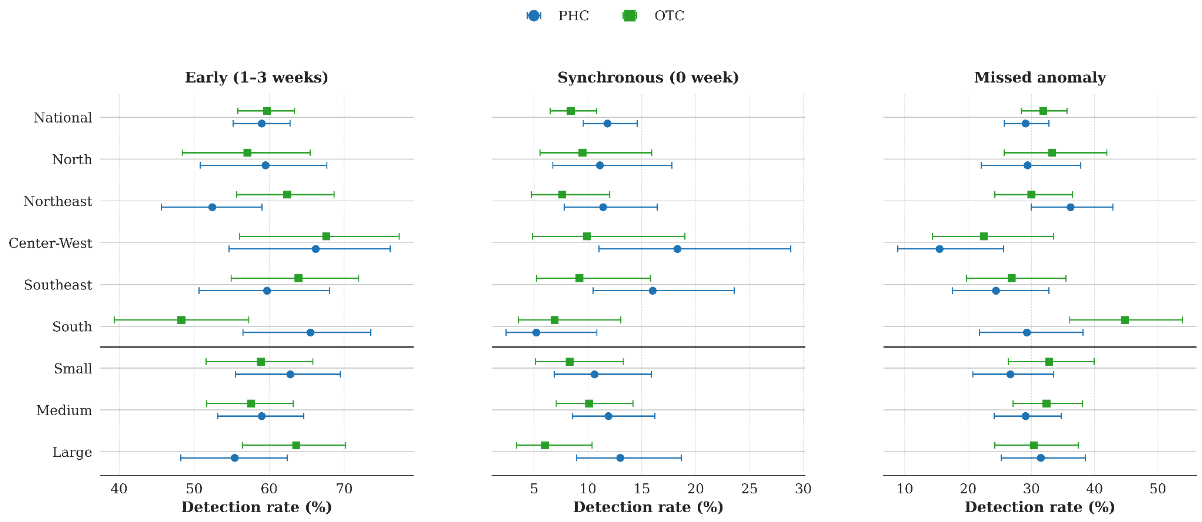


**Figure 2: Timeliness of early warnings detected in PHC and OTC time series relative to hospitalization surge events (as defined here) at national and regional levels.** This table summarizes the proportion of hospitalization surges that were anticipated (1 to 3 weeks early), detected in the same week (timely), or missed entirely based on PHC and OTC series. Results are presented for the national level, each geographic region, and by population size category .

Among regions, the PHC encounters have better performance in all regions, but Northeast region, when compared with the use of OTC drug sales data. Overall, specificity is also better in those regions for PHC data if compared with OTC drug sales (Figure 3).

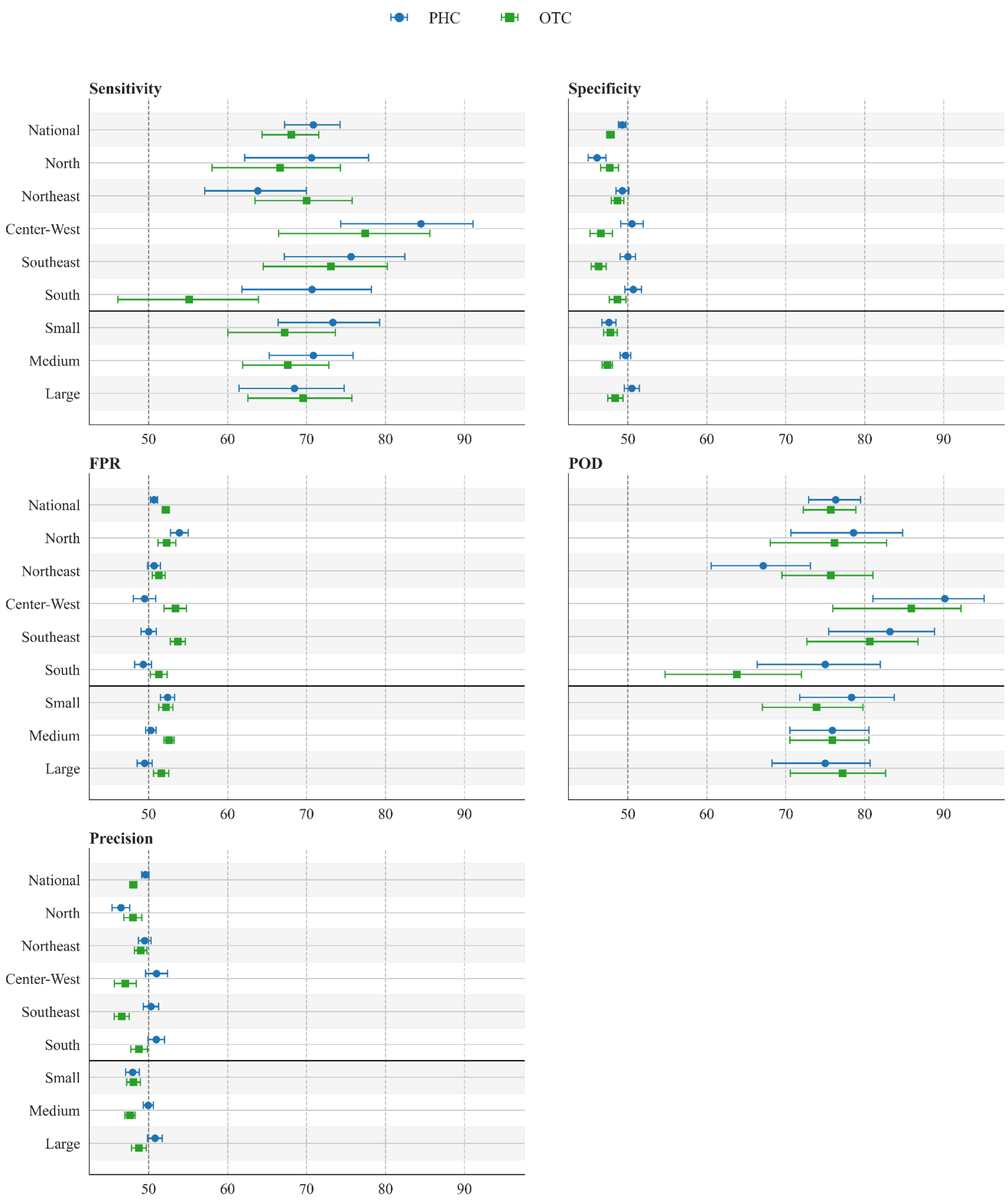


**Figure 3: Performance metrics for PHC encounters and OTC drug sales data in detecting hospitalization warnings at national, regional, and population size levels.** The figure presents the metrics of performance of Sensitivity, Specificity, Probability of Detection (POD), False Positive Rate (FPR), and Precision (PR) for both PHC and OTC time series across different geographic and demographic contexts.

Regarding population size, small-sized cities achieved the best sensitivity performance for PHC and large-sized cities had better performances for OTC (Figure 3). In contrast, PHC data shows lower performance in regions with larger population sizes, while lower performance is shown in regions with small populations for OTC data.

#### Supplementary Note 3

Model diagnostics

The following figures show the AIC distribution for the best NegBi-SPC model selected for each immediate region and each stream used.

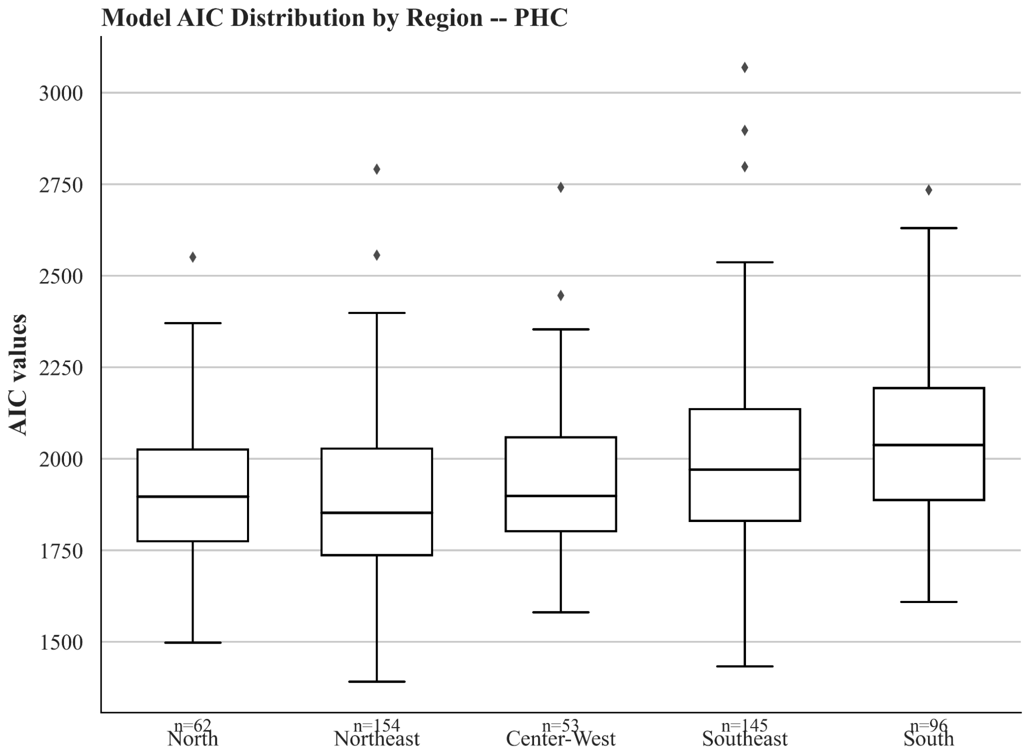


**Figure 1:** AIC distribution by macro-region category (North, Northeast, Center-West, South) corresponding to the best NegBi-SPC model adjusted to the PHC time series for each immediate region within these categories.

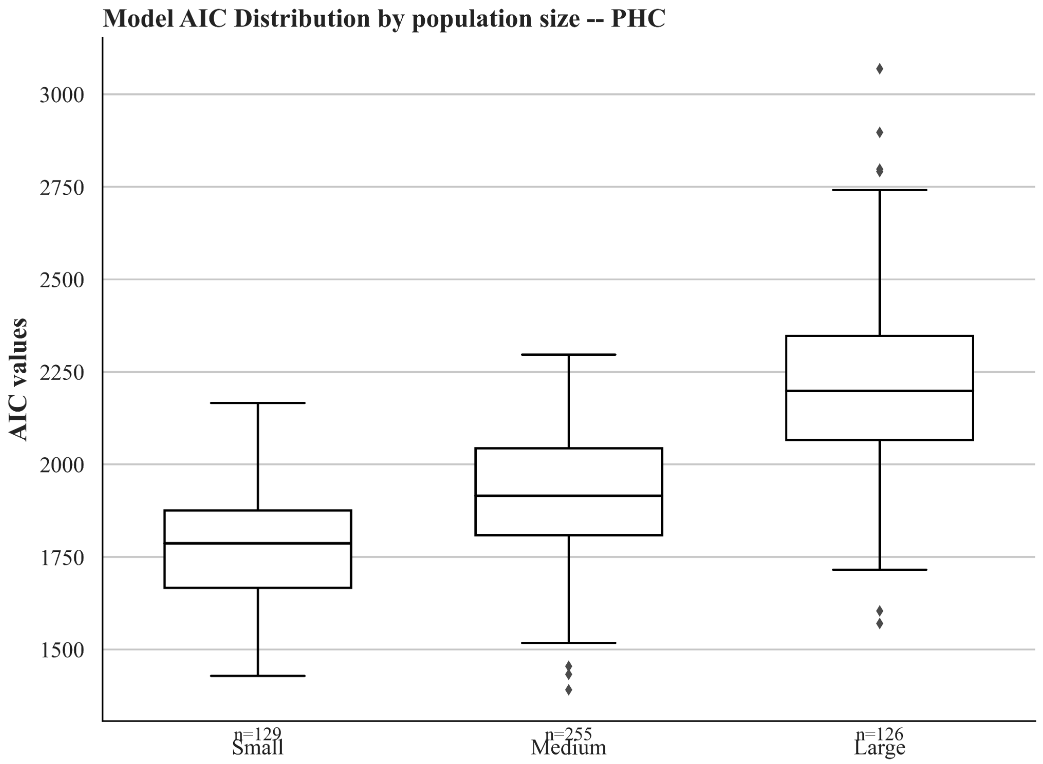


**Figure 2:** AIC distribution by population size category (Small, Medium, and Large), corresponding to the best NegBi-SPC models adjusted to the PHC time series for each immediate region within these categories.

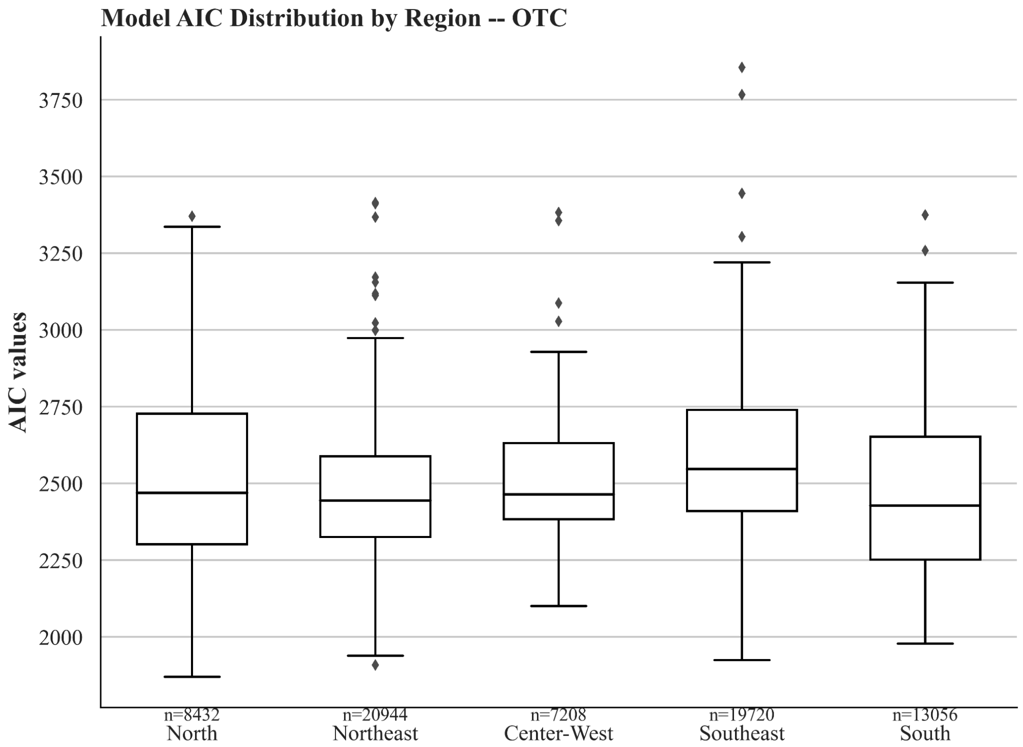


**Figure 3:** AIC distribution by macro-region category (North, Northeast, Center-West, South) corresponding to the best NegBi-SPC model adjusted to the OTC time series for each immediate region within these categories.

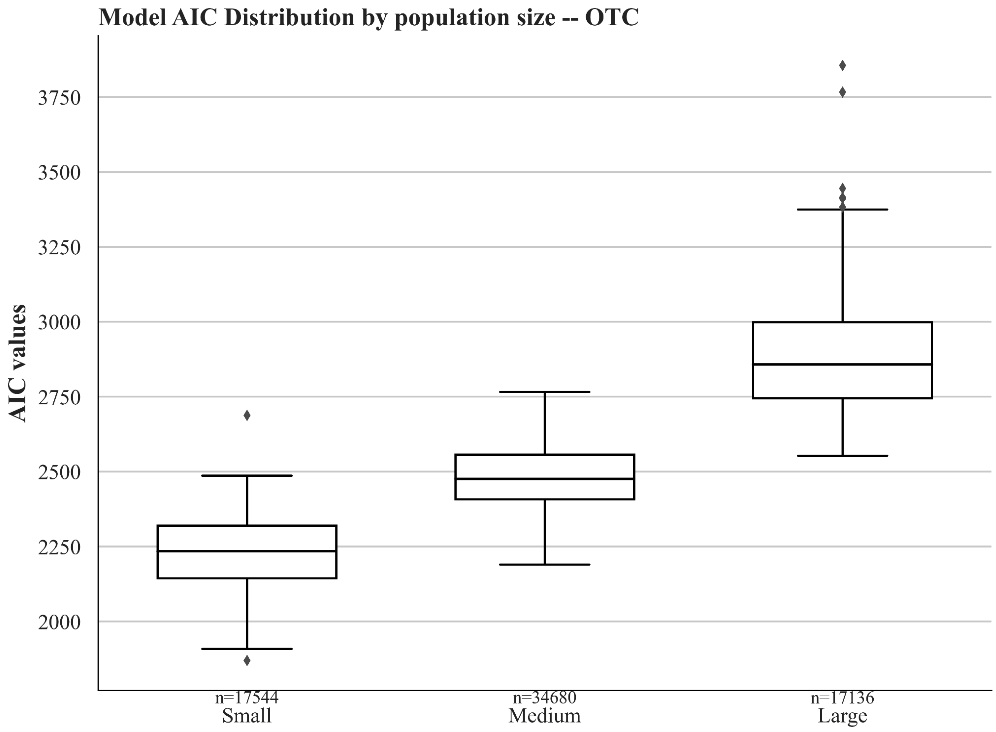


**Figure 4:** AIC distribution by population size category (Small, Medium, and Large), corresponding to the best NegBi-SPC models adjusted to the OTC time series for each immediate region within these categories.

Supplementary files 1 (PHC_ModelOutputs.csv) and 2 (OTC_ModelOutputs.csv) include variables supporting the evaluation of the NegBi-SPC model results, as detailed in Table 1 below.

Table 1: Variables supporting the evaluation of the NegBi-SPC model results.

| New column name | Description |
|---|---|
| State | Federative Unit (state) name where the immediate region is located. |
| ImmediateRegionCode | Unique IBGE code identifying the immediate geographic region. |
| ImmediateRegionName | Name of the immediate geographic region (IBGE classification). |
| EpiWeek | Epidemiological week of observation, formatted as YYYY-WW. |
| BestModelFormula | Model formula selected based on the lowest Akaike Information Criterion (AIC) value. |
| BestAIC | Minimum AIC value for the fitted model. |
| FittedValues_* | Fitted (predicted) values from the stream-only |

| | |
|---|---|
| | model. |
| Residuals_* | Model residuals (observed − fitted) for the stream-only model. |
| UpperControlLimit_* | Upper control limit (threshold for unusually high values). |
| LowerControlLimit_* | Lower control limit (threshold for unusually low values). |
| AnomalyAbove_* | Binary indicator: 1 if the observed value exceeds UCL. That variable defines the warning provided by the stream. |
| AnomalyBelow_* | Binary indicator: 1 if the observed value is below LCL. |
| Population | Estimated total population of the immediate region. |
| MacroRegion | Geographic macro-region of Brazil (North, Northeast, Center-West, Southeast, South). |
| PopulationCategory | Population size category (e.g., Small, Medium, Large), based on our predefined cutoffs. |
| * corresponds to a data stream considered PHC or OTC. | |
